# Client Awareness of a Specialised STI Wellness Clinic in Eswatini: Contextualised Marketing for a Stigma-free Client-Centred STI Clinic

**DOI:** 10.64898/2026.05.04.26352402

**Authors:** Yves Mafulu, Victor Williams, Phumlile Ndlovu, Khetsiwe L. Maseko, Sandile Ndabezitha, Nkosana Ndlovu, Siphesihle Gwebu, Nomcebo Matsenjwa, Benedictus Deku, Ncedo Mhlanga, Nkululeko Dube

## Abstract

1.

**Introduction:** Sexually transmitted infections (STIs) remain a major global public health challenge, with a particularly high burden in Southern Africa. In Eswatini, the burden of STIs, including HIV, is alarming, necessitating effective prevention strategies. Understanding clients’ engagement with STI services and how they learn about and access these services is vital for effective public health communication and service delivery. AHF Eswatini has implemented multiple demand creation strategies to improve awareness and utilisation of Wellness Clinic services, but evidence remains limited on which approaches are most effective from the client perspective.

**Objective:** This study aims to assess clients’ awareness, preferences, and perceptions of marketing strategies used by the clinic to inform marketing, communication and engagement approaches. Specifically, the study will examine the main sources of awareness, preferred communication channels, perceived effectiveness of outreach activities, and the relationship between awareness and service utilisation. This manuscript reports the study protocol prior to initiation of participant recruitment and data collection.

**Methods:** A cross-sectional analytical study will be conducted from February 2026 to June 2026 at the LaMvelase STI Wellness Centre. Participants will include clients attending the STI Wellness Centre regardless of HIV status. Individuals aged 15 and above will be recruited through systematic sampling after every 5^th^ client interval. Data will be collected via structured questionnaires and medical records. Clients will be surveyed on how they learned about the clinic, service satisfaction, and preferred communication channels. Statistical analyses will include descriptive statistics, evaluation of marketing reach and effectiveness, and logistic regression to identify associations. The analysis will also compare awareness patterns across demographic and key population groups and explore alignment between client-reported awareness and the demand creation strategies implemented by AHF Eswatini.

**Results:** This protocol describes a study designed to generate evidence on client awareness, communication preferences, and the effectiveness of marketing strategies for STI service utilisation. Findings will inform public health strategies and educational programs aimed at reducing STI rates and improving sexual health outcomes in Eswatini. The study is also expected to identify high-impact communication channels for different client groups and generate operational recommendations for improving demand creation efficiency.

**Conclusion:** This research will contribute valuable data to guide interventions and health policies and to design more effective interventions and communication strategies in high STI prevalence settings, ultimately supporting efforts to mitigate the impact of STIs and HIV in Eswatini.

## 2. Introduction

### 2.1. Background

Sexually transmitted infections (STIs) remain a major global public health challenge, with more than one million new infections acquired daily worldwide. The burden of STIs is further compounded by persistent gaps in prevention, testing, and treatment services, particularly in resource-limited settings while antimicrobial resistance and asymptomatic infection continue to challenge STI control efforts (1,2). Untreated STIs can lead to serious health complications, including infertility, increased risk of HIV transmission, and complications during pregnancy(2,3). The Southern Africa region has one of the highest rates of STIs, and estimates indicate that approximately 70% of new HIV infections globally occur in sub-Saharan Africa (4). In Eswatini, the burden of STIs, including HIV/AIDS, remains alarmingly high. In a study assessing the acceptability of point-of-care testing (POCT) among sexually active adults aged 18-45 years, 22% were found to be infected(5). In another study among adolescents and young people living with HIV (AYLHIV) in Eswatini, 15.5% of people aged 15-24 years tested positive, and 25% among females aged 20-25 years(6). Ginidza et al. (2017) found a prevalence of 19.4% among females aged 15-49 years. This is particularly concerning given the country’s high HIV prevalence rate, which is one of the highest in the world, standing at approximately 24.8% (Eswatini Ministry of Health, 2023). Among adolescents and young people in low- and middle-income countries, STI care-seeking is often delayed or avoided because of confidentiality concerns, stigma, cost, clinic environment, and provider attitudes (8)

In response to this high burden, AIDS Healthcare Foundation (AHF) Eswatini has established specialised STI Wellness Clinics as part of an integrated service delivery model aimed at improving access to STI screening, treatment, and prevention services, particularly among high-risk populations. To further enhance service uptake, AHF Eswatini has implemented a range of demand creation strategies, including community outreach, peer educator mobilisation, facility-based sensitisation, campaign-based interventions, and partner notification approaches. These strategies are designed to increase awareness, reduce stigma, and promote utilisation of STI services. Despite the availability of these services, including dedicated STI wellness clinics such as the AHF LaMvelase STI Wellness Centre, utilisation of these services depends heavily on client awareness, accessibility, perceived confidentiality, and acceptability. However, STI services utilisation remains suboptimal in many settings, often due to persistent stigma, low awareness, misinformation, and insufficiently targeted communication strategies, particularly among key populations (9,10).

Health marketing and communication strategies play a critical role in addressing these barriers by influencing how clients become aware of services, how they perceive them, and whether they decide to seek care. These strategies include digital platforms (e.g., Facebook, WhatsApp, TikTok), traditional media (radio/TV), community outreach events, peer-to-peer communication, and facility-based referrals, as reflected in the supplementary marketing survey tool (See Supplementary file 1). The effectiveness of these communication channels varies depending on demographic and behavioural factors such as age, gender, level of education, socioeconomic status, and risk group (Noar, 2008; Evans et al., 2015). In the context of Eswatini, where social media use is rapidly increasing, digital health marketing presents an opportunity to enhance awareness and engagement with STI services; however, there is limited empirical evidence on which communication channels are most effective in reaching different population groups, particularly key populations such as sex workers, MSM, adolescents, and young adults. Social media and online networking platforms have increasingly been used for sexual health promotion because they offer interactive, low-cost channels for reaching populations that may not be easily reached through traditional health communication approaches (13,14)

However, while routine program data demonstrates increases in clinic utilisation following demand creation activities, it does not provide insight into which strategies generated awareness, which channels were most influential, or how clients perceived these interventions. This highlights a critical gap between program-level evidence on utilisation trends and client-level understanding of awareness, perception, and behavioural drivers. Furthermore, client awareness is not only about exposure to information but also includes recall, perception, trust, and behavioural response to marketing messages, which ultimately influence health-seeking behaviour and service utilization (12,15,16). Evidence from low- and middle-income settings suggests that adolescent- and youth-friendly sexual and reproductive health services can improve access and utilisation when services are confidential, acceptable, accessible, and supported by community demand-generation approaches (17) The same can likely be said for key populations or any other population groups accessing such services.

### 2.2. Justification for the study

In Eswatini, no study, to our knowledge, has been conducted to assess the marketing approach and clients’ preferred communication strategies to inform them about the STI clinic activities and services. This represents a critical gap, as effective STI control depends not only on service availability but also on demand generation and informed utilisation of services. Without understanding how clients become aware of services and which communication channels influence their decision to seek care, health programmes, especially the ones requiring confidentiality and privacy, and easily stigmatising, risk investing in inefficient or misaligned marketing strategies. The availability of a structured marketing feedback tool provides a unique opportunity to systematically assess sources of awareness, exposure to marketing materials, preferred communication platforms, participation in outreach activities, and perceived effectiveness of outreach efforts. While programmatic evaluations can assess trends in utilisation and service uptake, they do not capture client-level awareness, recall, trust, or preferences. This study, therefore, complements programmatic monitoring by generating client-centred evidence that can explain why certain demand creation strategies do or do not work.

### 2.3. Potential use/relevance of study findings

The findings of this study can guide public health strategies, help refine educational programs, and enhance intervention methods tailored to reduce STI prevalence and improve sexual health outcomes. Furthermore, it addresses the unique needs of Eswatini’s population, where STI rates and HIV prevalence are significant, thus contributing to more targeted and effective health policies, marketing and communication strategies. Specifically, findings from this study can inform evidence-based marketing and communication strategies, identify high-impact communication channels for different population groups, support client-centred service delivery models, improve efficiency of outreach investments, and strengthen the linkage between community awareness and facility utilisation.

## 3. Aim and Objectives

### 3.1. Study questions

1. Which marketing strategy is effective in creating awareness about STIs and increasing service utilisation among the target population?
2. Which communication channels are most frequently used and trusted by clients?
3. How do marketing strategies influence client decisions to access STI services?
4. Is there alignment between implemented demand creation strategies and client-reported awareness?

### 3.2. Aim

To assess client awareness, exposure, preferences, and perceptions of health marketing and demand creation strategies, and to examine their association with reported service utilisation at the LaMvelase STI Wellness Clinic in Eswatini.

### 3.3. Objectives

1. To assess clients’ awareness of LaMvelase STI Wellness clinic and its services.
2. To identify the main sources through which clients learn about the STI Wellness Clinic
3. To describe client exposure to, recall of, and perceptions of marketing and demand creation strategies.
4. To determine clients’ preferred and trusted communication channels and assess their satisfaction with clinic accessibility and services.
5. To examine associations between client characteristics, marketing exposure, awareness sources, and reported service utilisation.
6. To formulate recommendations for optimising STI clinic marketing and demand creation strategies.

## 4. Methodology

### 4.1. Study design

This study will use a cross-sectional analytical design to assess client awareness, exposure, and perceptions of health marketing and demand creation strategies at the STI Wellness Clinic. The design will allow for both descriptive analysis of awareness levels, communication channels, and client preferences, as well as analytical assessment of associations between awareness, marketing exposure, and reported service utilisation.

### 4.2. Study Status and Timeline

At the time of submission of this protocol, the study has not yet commenced, and no participant data collection or data analysis has been conducted. Participant recruitment is scheduled to begin in February 2026 and is expected to be completed by June 2026. Data collection will occur concurrently with recruitment and is expected to be completed by June 2026. Data cleaning and analysis are anticipated to be conducted between June and July 2026. Study results are expected to be available from August 2026 onwards.

### 4.3. Study setting

The study will be conducted at the LaMvelase STI Wellness Centre, which operates within the LaMvelase Centre of Excellence, a standalone facility specialising in HIV care, serving approximately 15,000 registered patients. The STI Wellness Centre focuses on STIs and provides STIs screening, diagnosis, and treatment, and offers prevention commodities such as male and female condoms, lubricants, PrEP, PEP, and referrals for voluntary medical male circumcision (VMMC). Psychosocial support is also available to help clients manage the emotional aspects of sexual health. Open to both HIV-positive and HIV-negative individuals, the Wellness Centre aims to serve sexually active people at higher risk for HIV and other STIs, including sex workers and other key populations. Demand creation activities implemented around the clinic and through AHF Eswatini-supported programming include facility-based sensitisation, community outreach, peer educator mobilisation, campaign-based activities, and partner notification approaches.

### 4.4. Study population and participants

The study population will consist of clients who visit the Wellness Clinic during the study period. Any client attending the clinic, aged 15 years and above, regardless of HIV status or reason for visit, will be recruited. The study will include both first-time and repeat clients, allowing comparison between new clients whose attendance may be strongly influenced by awareness pathways and returning clients who can provide information on retention, satisfaction, and continued engagement.

### 4.5. Sampling and Sample Size

A systematic sampling approach will be used to recruit participants, whereby every 5^th^ client attending the clinic during the study period will be invited to participate, based on the estimated daily client flow. Potential participants who meet the inclusion criteria will be informed about the research and offered an opportunity to participate. Information on the study will include the objectives, associated risks of participating, and possible benefits. Those who agree to participate will be required to sign an informed consent form before they can be included. For this study, the outcome of interest is the proportion of clients aware of the clinic via specific marketing channels. Therefore, the sample size is calculated using:

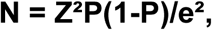

Where:

N = the required sample size

Z = the standard deviation at 95% confidence level (1.96).

P = Estimated prevalence of the outcome or proportion (expressed as a decimal). We will use P = 0.5 to ensure adequate power when the true awareness proportion is unknown, and as a standard for behavioural and awareness studies (18).

e =the standard error (0.05).

This gives **N = 384.** After adjusting for 10% non-response, the final sample size is **N = 422** participants. This sample size is considered adequate to provide stable estimates for proportions and to support subgroup and multivariable analyses of factors associated with awareness and service utilisation.

### 4.6. Inclusion and exclusion criteria

#### 4.6.1. Inclusion criteria

I. Clients aged 15 years and above who attend the Wellness Clinic will be eligible to participate, regardless of their HIV or PrEP status.
II. Clients who are willing and able to provide informed consent or assent, where applicable.

#### 4.6.2. Exclusion criteria

I. Clients less than 15 years of age.
II. Clients who were not attended at the Wellness STI clinic or did not receive any offered services.
III. Clients who are too ill, distressed, or otherwise unable to complete the interview at the time of recruitment.

### 4.7. Data management

#### 4.7.1. Data source, mining, and extraction

Data for the study will be obtained from an interviewer-administered survey. The survey is divided into sections – demographic characteristics, health information, sexual behaviour and health-related characteristics. The survey will be available in English and Siswati. Before the actual data collection, it will be piloted by the researchers on individuals who will not form part of the study participants. This is to ensure consistency and accuracy. The survey will be administered in a private room for confidentiality. Patient records available in the Client Management Information System (CMIS) and the Electronic Medical Record (EMR) will be the second source of data for the study. The questionnaire will specifically capture awareness sources, communication preferences, exposure to marketing materials, participation in outreach activities, preferred social media platforms, perceived effectiveness of campaigns, ease of accessing services, satisfaction with services, and suggestions for improvement. Where feasible, contextual information on implemented demand creation strategies and timing will also be reviewed to support the interpretation of findings, including information from the routine clients’ survey.

##### Study outcomes

The primary outcome is the awareness of the STI Wellness Clinic through specific communication channels. Secondary outcomes include exposure to marketing strategies, preferred communication channels, perceived effectiveness of outreach activities, service utilisation patterns, and client satisfaction.

##### Study Variables

The primary study variables will include demographic variables such as age, gender, sexual orientation, socioeconomic status and any other relevant demographic information. Additional study variables will include source of first awareness of the clinic, exposure to advertisements or marketing materials, preferred communication channels, participation in outreach activities, perceived effectiveness of outreach, clinic visit frequency, services accessed, ease of access, overall satisfaction, likelihood of recommending the clinic, and open-ended suggestions for service or outreach improvement.

#### 4.7.2. Data analysis plan

All data will be entered into an Excel spreadsheet to enable linkage between the different data sources. Data will be checked for completeness, accuracy and relevance. Data will be cleaned to remove any inconsistencies or incomplete records. Regular backups will be scheduled weekly, depending on the volume of data. Frequency tables and graphs will describe categorical variables and proportions. Data will be analysed using descriptive statistics to summarise frequencies and proportions of communication channel awareness, clinic satisfaction ratings, and service usage patterns. Comparative analysis by demographic subgroup (e.g., age, gender, key populations) may be conducted using chi-square tests. Findings will guide targeted outreach and communication strategies to improve clinic engagement and service uptake. Continuous variables will be presented using mean and standard deviation. To find whether there is a difference between groups or variables, a comparative analysis employing the T-test or Mann-Whitney test for continuous data and Pearson χ2 or Fisher’s exact test for categorical variables will be conducted. A p-value of <0.05 will be deemed statistically significant with a 95% confidence interval. Bivariate analysis will be conducted to explore associations between independent variables and key outcomes. Multivariable logistic regression analysis will be performed to identify independent predictors of clinic awareness and reported service utilisation, adjusting for potential confounders such as age, gender, and key population status. Stratified analysis may be conducted by age, gender, education level, and key population status. Where contextual program information is available, findings will be interpreted alongside implemented demand creation strategies to assess alignment or gaps between strategy implementation and client reach.

#### 4.7.3. Confidentiality and data safety

Data collected for this study will be anonymised after all the records have been linked before being made available to the research team. Completed questionnaires and other physical records will be kept in a lockable drawer. Only authorised people can access it. Electronic health records and data will be kept in secure servers with restricted access. Different levels of access will be assigned based on roles and responsibilities. All data will be anonymised to protect patient identities. Identifiers will be removed or masked in any dataset used for analysis or reporting. Backup copies will be stored in geographically separate secure locations to protect against data loss. All research team members will be trained in human subjects research, and confidentiality agreements will be required for all personnel handling the data.

### 4.8. Ethical considerations

#### 4.8.1. Ethical Review

Approval for the study has been obtained from the Eswatini Health and Human Research Review Board (EHHRRB) under Protocol Reference Number EHHRRB206/2025. The approval includes the current study as part of a broader approved research programme.

#### 4.8.2. Consent and participation

For participants aged 15–17 years, informed consent will be obtained directly from the participants in accordance with national guidelines on sexual and reproductive health (SRH) services, which allow adolescents to access SRH services independently. Given the sensitive nature of STI-related services and the potential risks associated with disclosure, parental or guardian consent will not be required. A waiver of parental consent will be sought from the EHHRRB, as requesting parental consent could compromise confidentiality and deter participation. All minor participants will provide written informed assent/consent, and appropriate safeguards will be implemented to ensure voluntary participation and protection of participants.

#### 4.8.3. Potential risk to participants

Participants face several potential risks. Privacy concerns include fears about the confidentiality of personal information and the clinic’s nature and location. There is a risk of data misuse. Additional concerns include social and relational impacts from disclosure (whether voluntary or accidental), challenges with understanding informed consent due to illiteracy, and potential coercion due to seeking health services. Given the focus on communication and awareness, some participants may also feel uncomfortable discussing how they first learned about the clinic or the perceived stigma associated with using STI services. Mitigation strategies will involve counselling and support to address emotional distress and secure data practices to ensure confidentiality. Transparent reporting and ethical reviews will prevent data misuse. Maintaining privacy and providing relationship support will help manage social risks. Clear communication and thorough explanations will aid informed consent, and emphasising the voluntary nature of participation will reduce coercion. These measures will ensure the ethical conduct of the research and the safeguarding of participants’ rights and well-being.

#### 4.8.4. Potential benefits to participants

Participants will benefit from all the services provided free of charge. Participants will gain valuable information about STI prevention, PrEP, and safe sexual practices, which can empower them to make informed decisions about their sexual health. They may also benefit from increased access to healthcare services, such as STI testing and counselling, leading to earlier detection and treatment of STIs and improved health outcomes. The research assistant will further explain that the study findings will add to the body of knowledge and contribute to improved service delivery by providing evidence-based approaches and marketing strategies. Findings may also contribute to more client-centred communication and outreach approaches that better match the needs and preferences of the populations served.

## 5. Funding

### 5.1. Funding source

This study is conducted using internal programmatic resources from AHF Eswatini. No external funding was received.

### 5.2. Budget

A summary implementation study budget is presented in the table below:

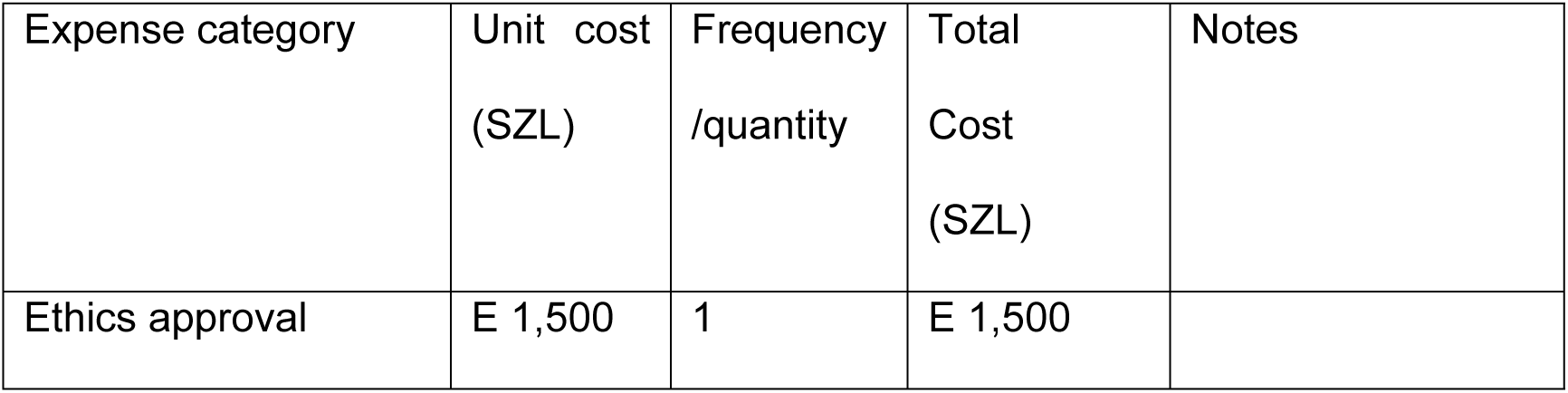
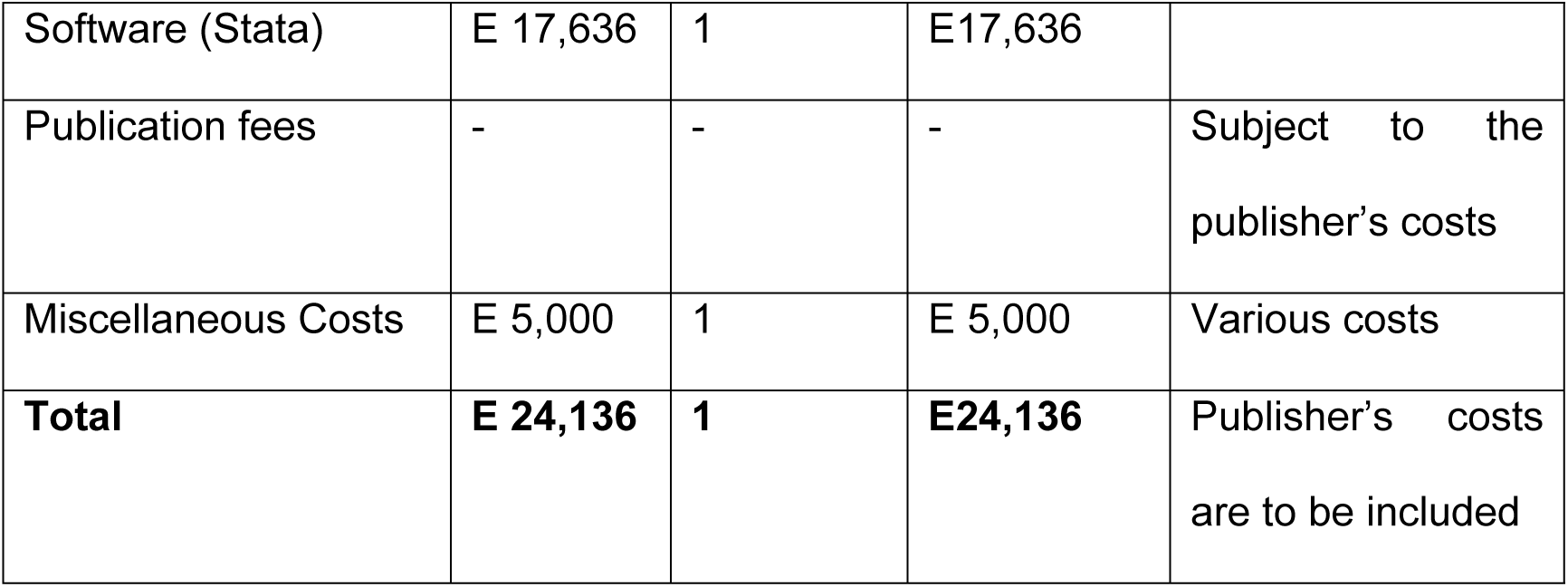

## 6. Timelines

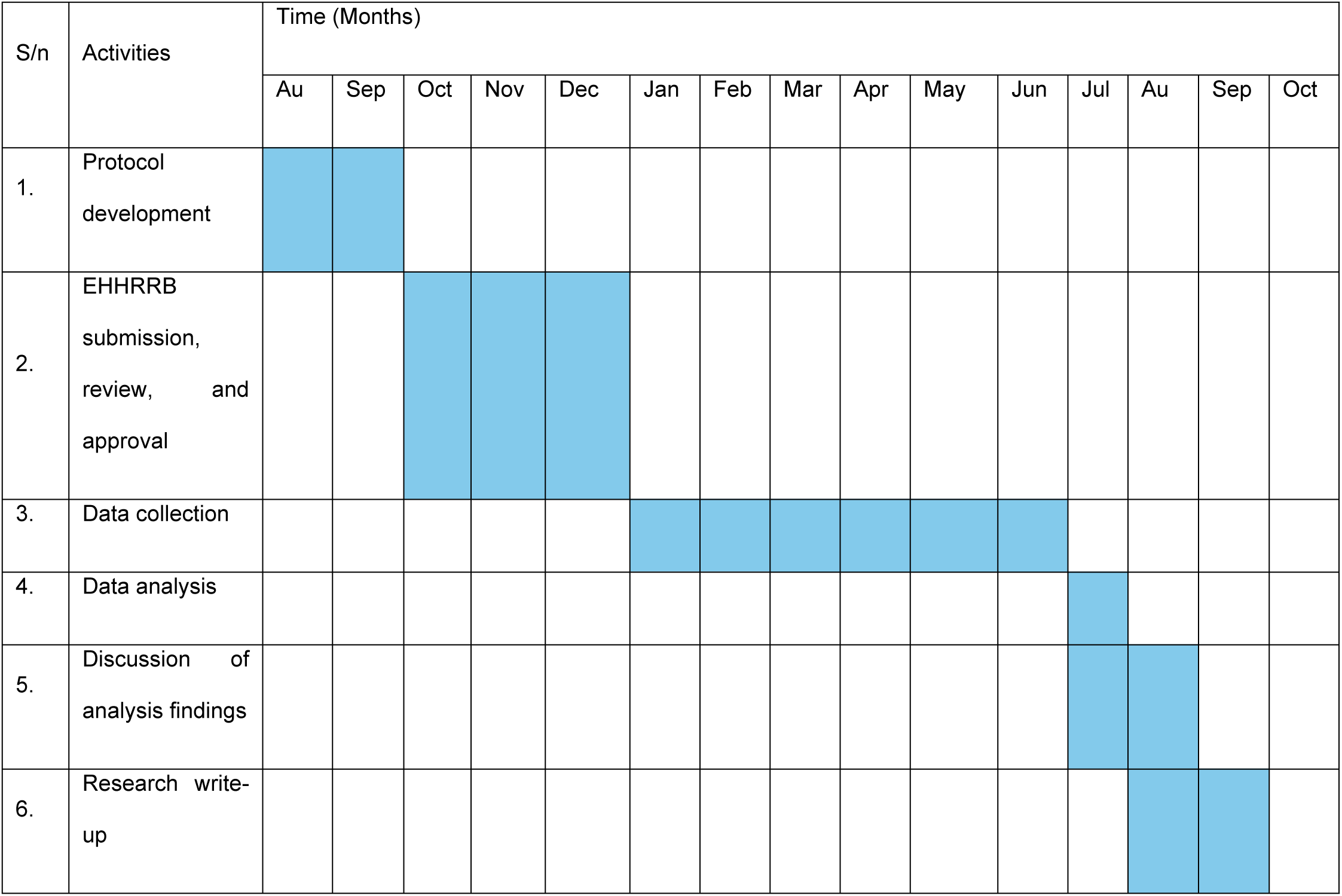
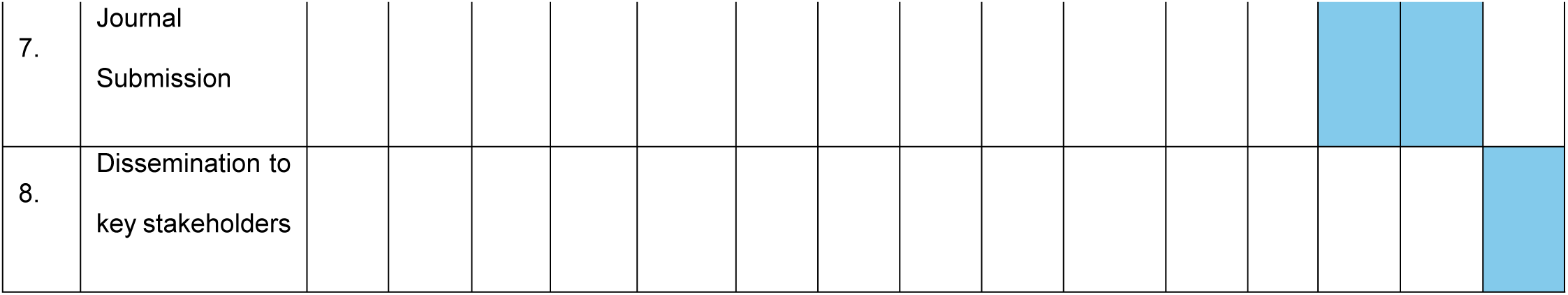

## 7. Discussion

### 7.1. Strengths

The study setting is a high-volume facility in the central part of the Manzini region and one of the two most densely populated administrative regions in Eswatini. Therefore, this could enhance the generalizability of the study findings. Moreover, the use of a specialised STI clinic as the study setting enhances the reliability of the data on communication and marketing strategies. The systematic sampling limits the potential for bias. The study also benefits from a structured client survey that can directly assess awareness pathways, perceptions of outreach, and preferred communication channels, thereby complementing routine program monitoring.

### 7.2. Limitations

The reliance on self-reported data might introduce social desirability biases. Moreover, this study will use a cross-sectional design; therefore, causality cannot be established since data will be collected at one point in time. This study will be facility-based; hence, the results of this study may not be generalisable to other settings or populations. The study may also be influenced by recall bias when participants report how they first learned about the clinic or whether they previously saw marketing materials. Furthermore, some demand creation activities may overlap in time, making attribution to a single strategy difficult. These limitations will be mitigated through careful questionnaire design, standardised interviewer training, and cautious interpretation of findings.

### 7.3. Dissemination of study findings

Through policy briefs, reports, meetings, and presentations, this study’s findings will be shared with key stakeholders, including national and international audiences. Abstracts will also be presented at national and international conferences to maximise the study’s impact and share evidence-based knowledge.

| **Audience** | **Method** | **Purpose** | **Timing** |
| --- | --- | --- | --- |
| **Ministry of Health (MOH)** | -Written study report<br>-Policy Briefs and Meetings | To inform policy updates and recommendations. | 2-3 months post-study |
| **AHF Africa Bureau and the global AHF Bureau** | - Reports<br>- Strategic Meetings<br>-PowerPoint presentation | - Discuss findings and implications.<br>-Update on impact and align global strategies.<br>Advocacy | 2-3 months post-study |
| **AHF Staff** | Internal Workshops/CMEs | Share findings and improve internal practices. | 1-2 months post-study |
| **HIV Care & Treatment TWG, Support partners** | -PowerPoint presentation<br>- Training, Sessions, Webinars,<br>- Collaborative Meetings | Share significant findings, review findings for integration into treatment guidelines | 3-4 months post-study |
| **Healthcare workers** | -PowerPoint presentation<br>-ReHSAR, NaHSAR, National conferences, Training Sessions, Webinars | Share significant findings and enhance understanding and application of findings. | 2-4 months post-study |

## 8. Data Availability Statement

Data generated or analysed during this study will be available from the corresponding author upon reasonable request.

## 10. Authors’ Contributions

YM conceptualised the study, developed the research protocol, and oversees implementation and ethical compliance. PN and KLM contributed to protocol writing, including review of study tools and consent materials. VW and SN reviewed the protocol, methodology, ensured compliance with research standards, and leads data management and analysis. NN provided input on study design and protocol refinement. SG, NM, BD, NMh, and ND contributed to protocol review, translation, and implementation support.

## 11. Acknowledgements

The authors would like to acknowledge the staff of the LaMvelase STI Wellness Clinic and all individuals who contributed to the development of this protocol.

The authors used artificial intelligence (AI) tools (ChatGPT, OpenAI) to assist with language refinement, editing, and structuring of the manuscript. All scientific content, study design, analysis, and interpretations were developed and verified by the authors. The authors take full responsibility for the accuracy and integrity of the work.

## Acronyms

AHF: AIDS Healthcare Foundation
ART: Antiretroviral Therapy
CMIS: Client Management Information System
CT: Chlamydia trachomatis
EHHRRB: Eswatini Health and Human Research Review Board
EMR: Electronic Medical Record
HIV: Human Immunodeficiency Virus
MOH: Ministry of Health
NG: Neisseria gonorrhoea
NaHSAR: National HIV Semi-Annual Review
PEP: Post-Exposure Prophylaxis
PI: Principal Investigator
PrEP: Pre-Exposure Prophylaxis
RPR: Rapid Plasma Reagin
SHIMS: Swaziland HIV Incidence Measurement Survey
SPSS: Statistical Package for the Social Sciences
STI: Sexually Transmitted Infection
STATA: Statistical Analysis Software
VMMC: Voluntary Medical Male Circumcision
WHO: World Health Organization

## Appendices/Supplementary files

- Supplementary file 1: Marketing strategy feedback Data collection tool (English and Siswati versions)
- Supplementary file 2: Consent form (English and Siswati versions)
- Supplementary file 3: Confidentiality and Non-disclosure Statement
- Supplementary file 4: Conflict of Interest Statement.

